# Assessing the risk of spread of COVID-19 to the Asia Pacific region

**DOI:** 10.1101/2020.04.09.20057257

**Authors:** Freya M. Shearer, James Walker, Nefel Tellioglu, James M. McCaw, Jodie McVernon, Andrew Black, Nic Geard

## Abstract

During the early stages of an emerging disease outbreak, governments are required to make critical decisions on how to respond appropriately, despite limited data being available to inform these decisions. Analytical risk assessment is a valuable approach to guide decision-making on travel restrictions and border measures during the early phase of an outbreak, when transmission is primarily contained within a source country. Here we introduce a modular framework for estimating the importation risk of an emerging disease when the direct travel route is restricted and the risk stems from indirect importation via intermediary countries. This was the situation for Australia in February 2020. The framework was specifically developed to assess the importation risk of COVID-19 into Australia during the early stages of the outbreak from late January to mid-February 2020. The dominant importation risk to Australia at the time of analysis was directly from China, as the only country reporting uncontained transmission. However, with travel restrictions from mainland China to Australia imposed from February 1, our framework was designed to consider the importation risk from China into Australia via potential intermediary countries in the Asia Pacific region. The framework was successfully used to contribute to the evidence base for decisions on border measures and case definitions in the Australian context during the early phase of COVID-19 emergence and is adaptable to other contexts for future outbreak response.

## Introduction

On December 29, 2019, Chinese authorities reported a cluster of cases of atypical pneumonia in the city of Wuhan, Hubei Province, later identified to be caused by a novel coronavirus, SARS-CoV-2 [1]. The disease caused by this virus is now known as Coronavirus disease 2019 or COVID-19 [2]. The number of confirmed cases and geographical extent of COVID-19 has increased significantly since December 2019. As of February 29, 2020, 79,394 cases had been confirmed in mainland China, including 2,838 deaths. A further 6,009 cases and 86 deaths had been reported outside of China — and more than half of these cases had been reported in the Asia Pacific region. Multiple countries outside of China had reported sustained local transmission [3].

When a novel pathogen such as COVID-19 emerges, governments are required to make critical decisions on how to respond appropriately, despite limited data being available to inform these decisions. The extent of transmission of COVID-19 in mainland China changed rapidly during the first month of the epidemic. From January 22, 2020 Chinese authorities progressively implemented strict mobility restrictions on the people of Wuhan, and on travel out of the affected area to other parts of China. Internationally, border screening measures were implemented, and travel restrictions were imposed by both governments and airlines [4].

For example, Australian authorities placed restrictions on all travel to Australia from mainland China on February 1, in order to reduce the risk of importation of the virus. Only Australian citizens and residents (and their dependants) were permitted to travel from China to Australia [5]. These restrictions remained in place at the time of writing.

The day before Australia imposed these restrictions (January 31), 9,720 cases of COVID-19 had been reported in mainland China. Australia had detected and managed 9 imported cases, all with recent travel history from or a direct epidemiological link to Wuhan [6, 7]. A further 120 cases had been confirmed outside of China, including 95 cases in countries of the South East Asia and Western Pacific regions [7].

At that time (January 31), models estimated that 75,815 individuals (95% CrI 37,304–130,330) had been infected in Greater Wuhan up to January 25 (with WHO reporting 1,320 confirmed cases for the same time period) and projected that the epidemic could peak in Wuhan as early as late February [8]. Before the restrictions, Australia was expecting to receive approximately 200,000 air passengers from mainland China during February 2020 [9]. Travel numbers fell dramatically following the imposed travel restrictions.

Prior models suggest that travel restrictions are unable to completely prevent the international transmission of viral respiratory illnesses [10, 11]. Screening methods are unable to identify all cases, due to limitations in the sensitivity and specificity. Exposed travellers can complete a journey during the incubation period when they are undetectable, and some exposed travellers may never show symptoms at all [10, 11]. Furthermore, when applied early in an epidemic, screening methods that rely on detecting fever will identify many more cases of other respiratory diseases such as influenza than the target disease. While travel restrictions are highly unlikely to prevent the ultimate importation of COVID-19 [12], they can reasonably be anticipated to delay the establishment of an epidemic in a country, buying valuable time for health authorities to establish response measures.

Analytical risk assessment is a valuable approach to guide decision-making on travel restrictions and border measures during the early phase of an outbreak [13, 14, 15]. While importation risk into a country of interest from a source country is a general problem for emerging diseases, the specifics of risk assessment should consider the status of both the evolving epidemic and the local and global public health response.

Here, we consider the COVID-19 importation risk to Australia from countries other than China at a time when China was the only country reporting uncontained local transmission and limited cases had been reported in other countries. With air-travel restricted from China, and strict quarantine measures in place for those allowed to return, a key concern for Australia was the epidemic status of other countries with large (and unrestricted) travel volumes to Australia.

We introduce a modular framework for assessing the risk of COVID-19 being imported from a source country (here China) to a country of interest (here Australia) via other intermediary countries in the region. We have focused on the risk of (potentially undetected) spread to countries in the South East Asia and Western Pacific regions because they are highly connected to both China and Australia, relative to the rest of the world. Our analysis takes into account the COVID-19 epidemiological situation and mobility restrictions imposed as of mid-February but is adaptable to the analyses required during later phases of the outbreak.

The framework was developed to provide a rational basis for decision-making on border measures and case definitions in Australia at a time when global transmission of COVID-19 was not yet established, which was no longer the case at the time of writing. While the detailed analysis presented here is specific to Australia and the South East Asia and Western Pacific regions during the early phase of COVID-19 emergence, the framework itself is adaptable to other contexts for future outbreak response.

## Methods

### Overview

A framework was developed to assess the risk of COVID-19 infections being imported by passengers travelling on flights from the South East Asia and Western Pacific regions to Australia as of February 19, 2020.

The framework includes a series of analyses based on current epidemiological evidence, patterns of air travel, and model components described in De Salazar and colleagues (2020) [12] and developed by the authors. Our framework considered a single point of origin for all exported infections, as China was the only country reporting uncontained transmission at the time of analysis. Each step of the analysis is outlined in Figure 1 and described in more detail below.

**Figure 1:**
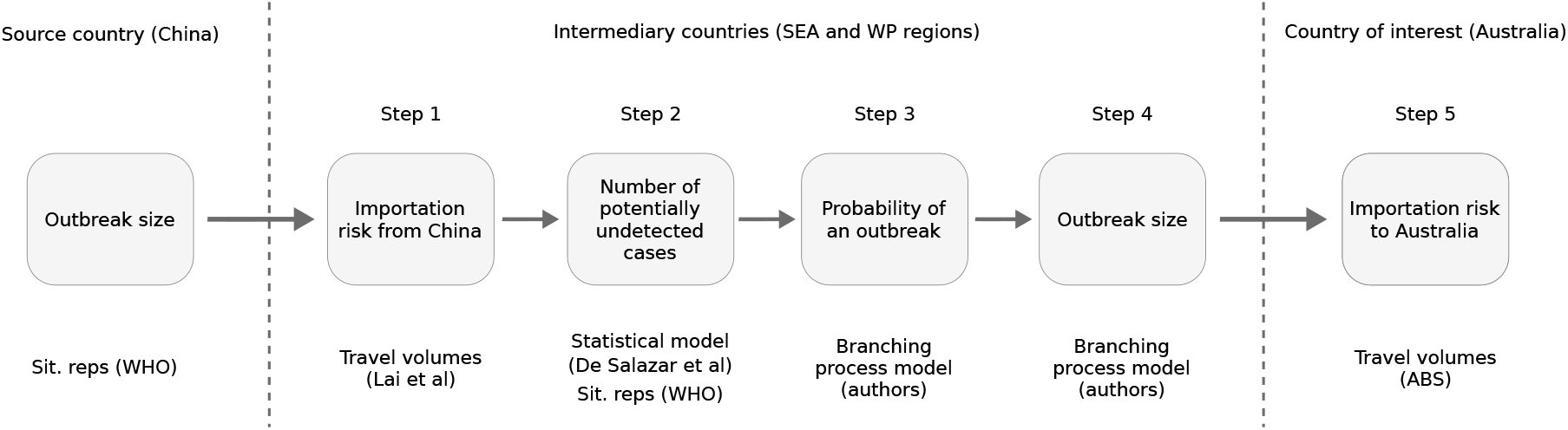
Key quantity estimated at each step of the full analysis of COVID-19 importation risk to Australia (country of interest) via countries in the South East Asia and Western Pacific regions (intermediary countries), given travel restrictions from mainland China (source country) (top). A brief indication of the relevant data and/or method used at each step (bottom).

### Step 1 – Importation risk from China to intermediary countries

SARS-CoV-2 first emerged in China [1] and hence the risk of importation for countries in the South East Asia and Pacific regions, in the early stages of the outbreak, was primarily dependent on travel from China. The expected numbers of imported cases in each intermediary country was estimated using an approach proposed by De Salazar and colleagues (2020) based on air travel volume estimates from China since COVID-19 emergence [12]. Their model estimates the expected number of imported cases in countries by regressing the number of imported cases reported by each country against their relative incoming travel volumes from China (under unrestricted travel). They assume that the expected case count would be linearly proportional to air travel volume. Bootstrap sampling was used to estimate 95% confidence intervals. We fitted the model to reported cumulative case counts for each country extracted from WHO situation reports 2, 9, 16, 23, and 30 (i.e., one per week from January 22).

### Step 2 – Number of potentially undetected cases in intermediary countries

The number of potentially undetected introductions in each intermediary country was based on the discrepancy between expected (Step 1) and reported cases (noting that cases due to local transmission were excluded from these counts). Under the assumption that all reported cases were effectively isolated (24 hours after symptom onset) with reduced risk to onward transmission, the difference between the expected and reported numbers of cases per country provided a crude estimate of the number of unreported cases. We assumed unreported cases were undetected and therefore more likely to contribute to local transmission and potentially a large outbreak.

### Step 3 – Probability of an outbreak in intermediary countries

A branching process model was used to generate stochastic projections of the initial stages of an outbreak for each country. This model incorporates country-specific rates of COVID-19 importation (as estimated in Step 1) and country-specific detection probabilities based on the ratio of reported cases to expected cases (with a maximum detection probability of 1). We assumed an R0 of 2.68 (within the range estimated for COVID-19 in Wuhan in early January [8]), no individual-level variation in transmission and independence of all undetected introductions [13]. The probability of local transmission was defined as the proportion of simulations with no locally transmitted cases 5 weeks after simulation commenced (i.e., February 26). Details of this branching process model are provided in the Supplement.

### Step 4 – Estimated size of an uncontained outbreak in intermediary countries

The stochastic transmission model described in Step 3 was also used to estimate the likely number of locally transmitted cases in each intermediary country, conditional on local transmission occurring. This model assumed no public health intervention, and that both importation rate and detection probability were constant over time. The transmission model was run from January 22, with epidemic curves (separated by imports and local transmission) projected forward by one week beyond the last data collection date (February 26).

### Step 5 – Importation risk from intermediary countries to a country of interest

The likelihood that a passenger arriving in Australia from a country in the South East Asia or Western Pacific region would be infected by COVID-19 will depend on the prevalence in that country, and the travel volume from there to Australia.

Given the wide confidence intervals on estimated prevalence, and the multiple stochastic factors, there is considerable uncertainty involved in such estimates. For the purpose of this initial risk assessment exercise, we simply provided the prevalence estimates and travel volumes. Travel volumes from countries within the South East Asia and Western Pacific regions to Australia were extracted from the Australian Bureau of Statistics [9, 16].

## Results

### Assessing the overall risk of case importation into a country of interest from intermediary countries

We present a ranked table of aggregate risk of COVID-19 infections being imported into the country of interest (Australia), based on sustained transmission occurring in the source country (China) by originating country of travel (Figure 2). The table includes key quantities estimated at Steps 1 to 5 of the analysis outlined in the methods and depicted in Figure 1. The potential number of undetected imported cases (as of February 19, 2020) in each country of the South East Asia and Western Pacific regions (intermediary countries) is displayed in Figure 3.

**Figure 2:**
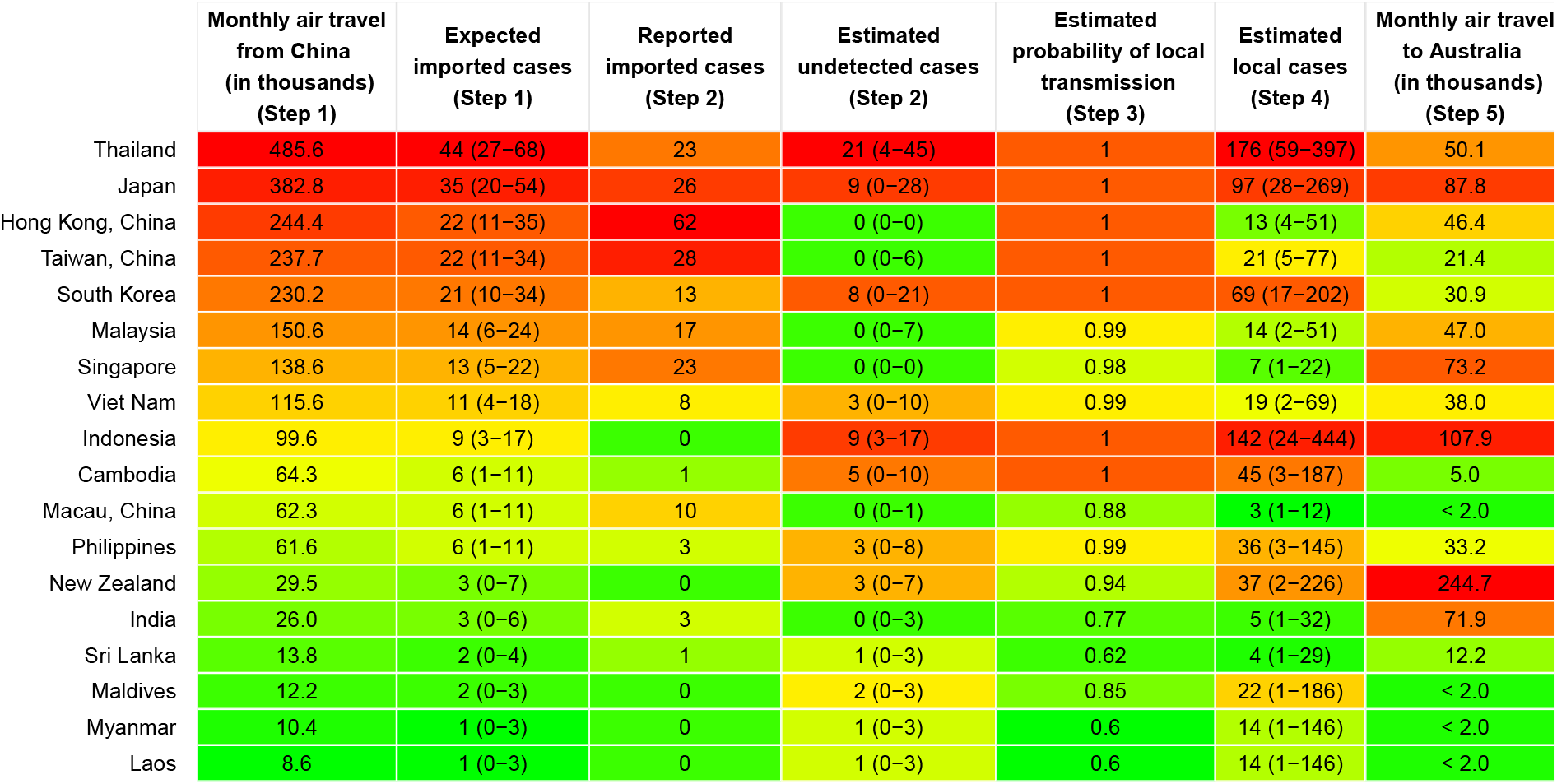
Summary table of indicators for assessing importation risk of COVID-19 to Australia from countries in the South East Asia and Western Pacific regions. Analysis as of February 19. At this time, the only country reporting uncontained/sustained transmission was China and restrictions to travellers from mainland China to Australia were imposed. Relative risks are indicated by colour shading, with red representing the highest risk and green the lowest.

**Figure 3:**
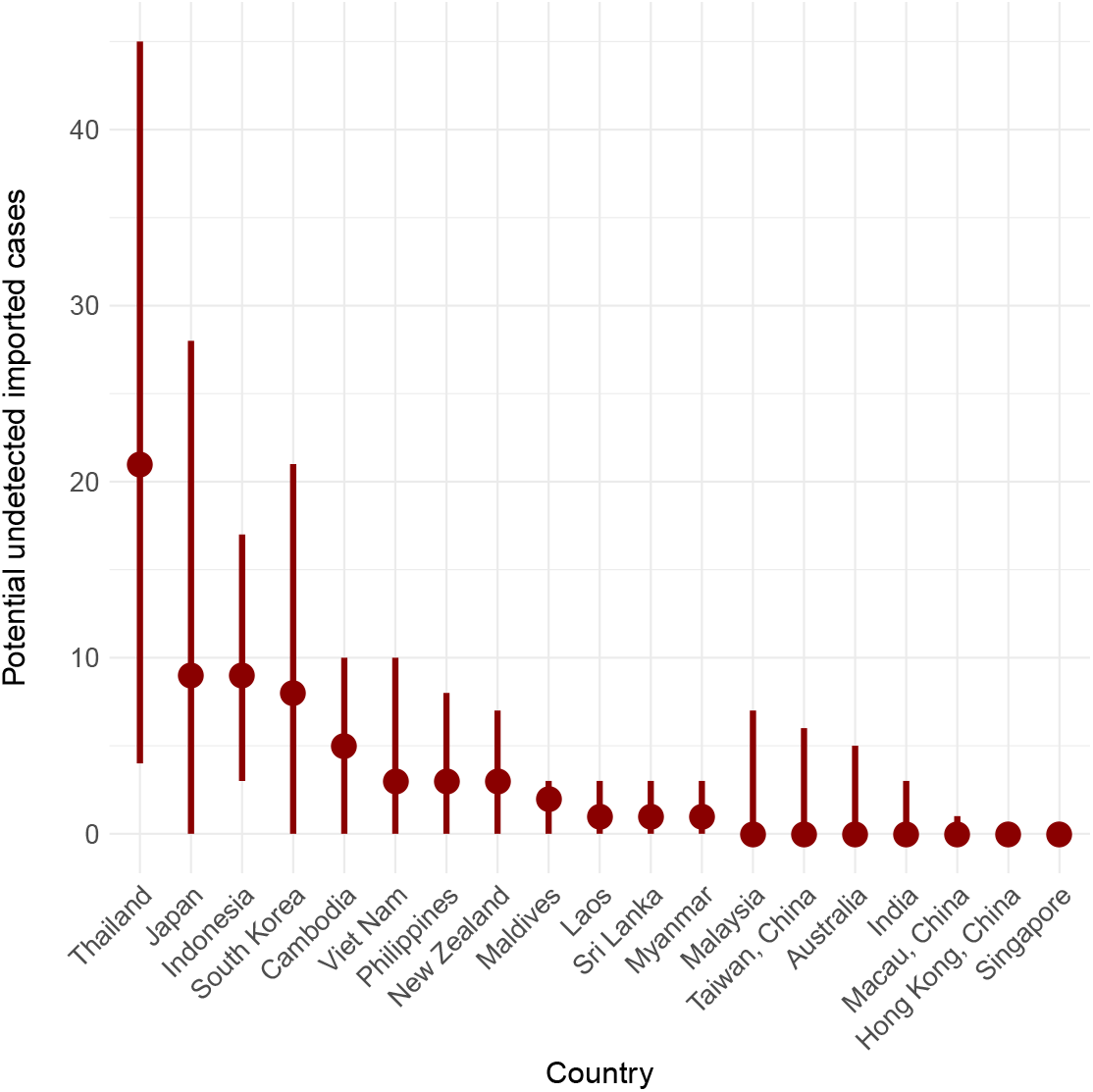
Estimated number of undetected cases and 95% confidence intervals for each country in the South East Asia and Western Pacific regions as of February 19, 2020.

Projected epidemic curves for selected countries are shown in Figure 4 (curves for all countries considered are provided in the Appendix), with reported imported and local cases also shown for context. All four countries shown have high levels of expected imported cases (Figure 2). Thailand and Indonesia have fewer reported cases than expected, and hence a higher estimated number of undetected cases (Figure 3), leading to projection of considerable undetected local transmission. In contrast, Malaysia and Singapore both reported a number of cases equal to or greater than expected, and hence have a lower estimated number of undetected cases, leading to projection of more modest levels of local transmission. Note that the projected levels of transmission in Singapore are lower than the actual reported cases. These known local cases were not further incorporated into our analysis, but rather signalled to decision makers that the likelihood of further undetected transmission was relatively low.

**Figure 4:**
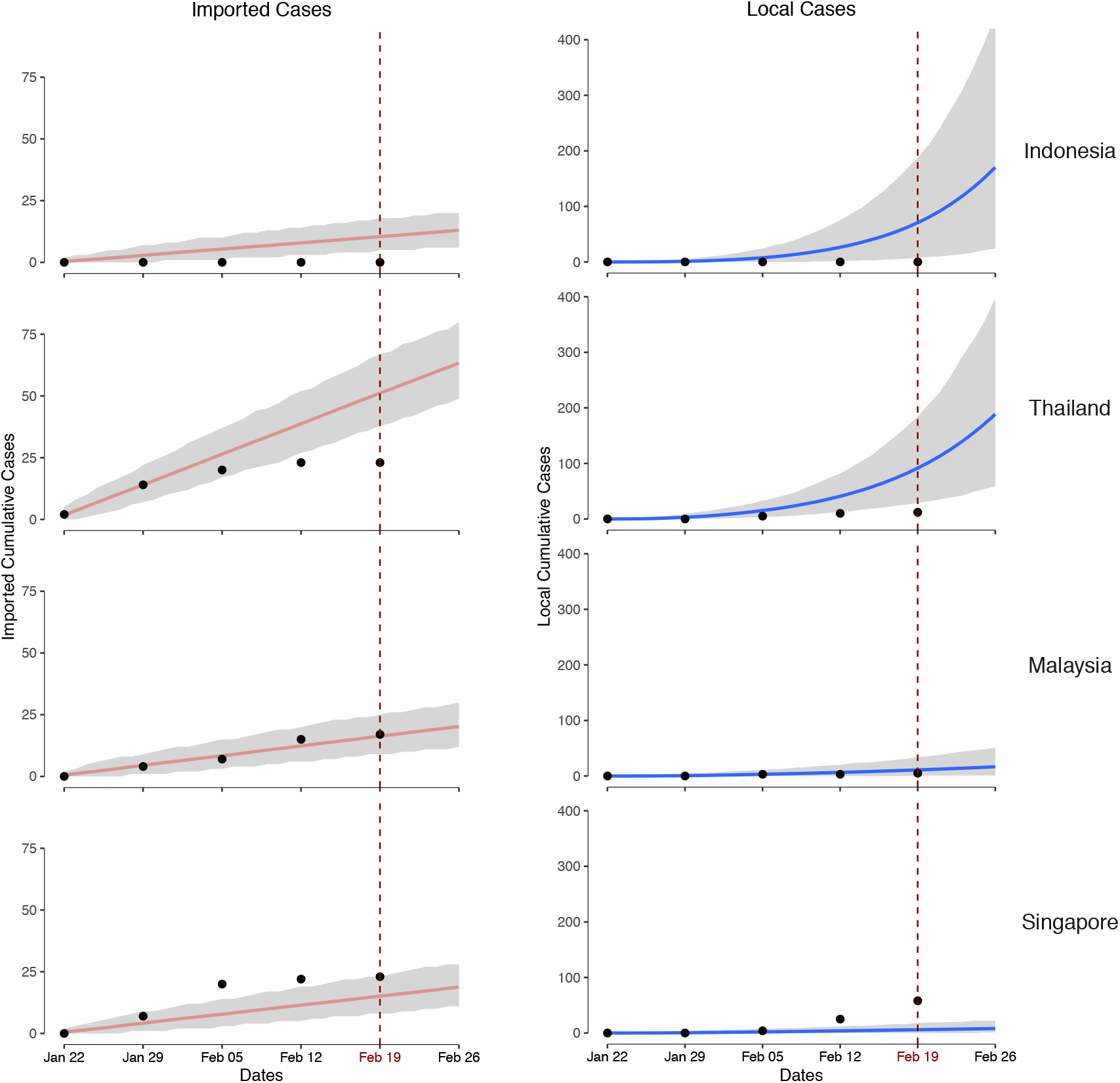
Imported cases (left panels) and epidemic curves (right panels) from January 22 using WHO data up to February 24, and projecting forward to March 4, for selected countries. Lines and shaded regions in each panel show median and 95% quantiles for the cumulative number of cases (imported or locally transmitted). Black points show cumulative imported cases (left panels) and cumulative local cases (right panels) for each country, as reported by WHO.

## Discussion

We developed a framework to assess the importation risk of COVID-19 into Australia during the early phase of the epidemic, from late January to mid-February 2020. The dominant importation risk to Australia at the time of analysis was directly from China, as the only country reporting uncontained transmission. With travel restrictions from mainland China to Australia imposed from February 1, our framework was designed to consider the importation risk from China (source country) into Australia (country of interest) via potential intermediary countries. We focused on countries in the South East Asia and Western Pacific regions as potential intermediary countries since they are highly connected to both China and Australia, relative to the rest of the world.

Intuitively, intermediary countries that pose the highest overall risk of importation to a country of interest are those with high connectivity to both the source country and country of interest. High connectivity to China increases the probability of an outbreak of COVID-19 in an intermediate country, and consequently, someone resident in that intermediate country and travelling to the country of interest has a higher likelihood of being exposed.

The risk table (Figure 2) was developed for two purposes. First, to provide evidence of likely exposure over the preceding week to inform the epidemiological case definition, tailored to the source country and for use in near patient decision making for practitioners in the country of interest (here Australia) caring for returned travellers. Second, projection of the likely future epidemic course helped to inform travel advisories and the likely utility of border restrictions.

The framework we have developed provides a risk assessment from solely from an epidemiological perspective. It does not consider the potential social, political and economic implications of future border measures and mobility restrictions which are both substantial [17] and will exert an influence on epidemiology as people change their behaviour [18]. This framework is therefore only one key element to be considered by decision-makers contemplating possible border policies and mobility restrictions, which are ultimately a political determination.

A significant strength of our approach is in its transparency with respect to both individual factors that contribute to overall risk and the relationship between model projections and the reported epidemiology. This enables decision-makers to incorporate their own expertise when interpreting the outputs. Even if limited or highly uncertain data is available to inform absolute estimates of risk associated with plausible importation routes, comparisons of relative risk using our approach are still possible and valuable. In addition, each analysis component has modest data requirements and low computation cost, making rapid preliminary assessment across a range of countries feasible. As the outbreak progressed, mismatches between model projections and observed epidemiology could be readily observed and incorporated into decision making.

A strength of our modular approach is that it enables individual components to be adapted as new data and models become available, or as information needs change in response to the evolving situation. The breakdown of our workflow (Figure 1) provides clear guidance on how to adjust the method. Importantly, the framework itself still applies, but components of the analysis would need to be adapted.

As implemented, our estimates of importation rates into countries only consider air travel (Step 1). Several countries/regions have high volumes of land travel, for example mainland China and Hong Kong. Not accounting for these would lead to an underestimate of importation risk to these countries/regions. Furthermore, the estimated importation rates are relative to global reports of imported cases of COVID-19. If there was systematic under-detection of COVID-19 across all countries at the time of analysis [19], this would lead to a systematic underestimation of importation rates. Our approach was not able to estimate rates of import prior to January 22nd when case counts for countries started being reported by the WHO.

Our analysis framework is specific to a scenario in which a dominant source of infection is mitigated by border measures. Once established outbreaks occur in countries other than the primary source, assessing importation risk becomes more complex. Further to this, we primarily focused on identifying *undetected* epidemics — over the course of time emerging *observed* data from many countries became more influential in the risk appraisal. Thus, as it stands, our analysis is only applicable in the early stages of an outbreak, a feature shared with many other models of global infection links.

In detail, by the time of writing, substantial transmission has been reported in a number of additional countries, in particular Iran, Italy and South Korea [3]. In order for our tool’s risk assessment to reflect the true risk of importation to the country of interest, components of the analysis, specifically Steps 1 and 2, would need to be adapted to account for high levels of known local transmission in countries other than the source country.

Returning to the exemplar study that motivated the development of our framework (risk of importation of COVID-19 into Australia), should a large outbreak occur in one or more countries in the South East Asia and Western Pacific regions, additional intra-regional transport connections would be associated with importation risk to countries in the region. More detailed modelling that incorporates intra-regional travel and information on known local transmission could then be included (at Steps 1 and 2) and updated to reflect emerging epidemic intelligence. This would allow the framework to be applied during such a phase of the outbreak.

A further strength of our approach is its use of a stochastic model, including control efforts, for early epidemic response in intermediate countries. Here, without access to detailed additional information on intermediate country’s capacity to respond, we used a simple model of country response capacity. As outbreaks progress, differences in how effectively countries are able to respond, both in terms of their health system capacity, and their ability to implement population-based measures such as social distancing may introduce systematic effects that are not captured by the framework. With additional information or dedicated further research in anticipation of future global events, this additional information could be incorporated into the framework (at Steps 3 and 4) to improve predictive capabilities. It is also worth noting that here we used a stringent definition of outbreak control (truly no cases) compared to other approaches in the literature, which may define control as a substantially smaller number of cases compared to baseline [20].

In conclusion, by developing a modular framework that describes not only the underlying mathematical models of transmission and control, but how each component integrates with the next to generate an overall assessment of importation risk of an emerging disease, we have provided a decision-making tool that is flexible to the analysis requirements at different phases of an outbreak. The framework provides an evidence base for decisions on border measures and case definitions, and it has been successfully used during the early phase of the COVID-19 response in the Australian context, when limited cases had been reported outside of mainland China.

## Data Availability

All data used is available from existing preprints/publications and the Australian Bureau of Statistics website.

## Notes

### Competing Interest Statement

The authors have declared no competing interest.

### Funding Statement

This material is based on work funded in part by the Australian Department of Health. JMcV is supported by a NHMRC Principal Research Fellowship.

